# Causal associations between female reproductive behaviors and psychiatric disorders: a lifecourse Mendelian randomization study

**DOI:** 10.1101/2023.03.20.23287480

**Authors:** Yifan Yu, Lei Hou, Yutong Wu, Yuanyuan Yu, Xinhui Liu, Sijia Wu, Yina He, Yilei Ge, Yun Wei, Fengtong Qian, Qingxin Luo, Yue Feng, Xiaojing Cheng, Tiangui Yu, Hongkai Li, Fuzhong Xue

**Author notes:** **Corresponding author:** 1. Fuzhong Xue, Address: School of public health, Shandong University, 44 Wenhua, West Road, Jinan, Shandong province, China, 2. Hongkai Li, Address: School of public health, Shandong University, 44 Wenhua West Road, Jinan, Shandong province, China, 3. Tiangui Yu, Address: Shandong Mental Health Center, Jinan, Shandong province, China. These authors contributed equally to this work (Yifan Yu, Lei Hou). These authors also contributed equally to this work (Tiangui Yu, Hongkai Li, Fuzhong Xue).

## Abstract

**Background:** The timings of reproductive life events have been examined to be associated with various psychiatric disorders. However, studies have not considered the causal pathways from reproductive behaviors to different psychiatric disorders. This study aimed to investigate the nature of the relationships between five reproductive behaviors and twelve psychiatric disorders.

**Methods:** Firstly, we calculated genetic correlations between reproductive factors and psychiatric disorders. Then two-sample Mendelian randomization (MR) was conducted to estimate the causal associations among five reproductive behaviors, and these reproductive behaviors on twelve psychiatric disorders, using genome-wide association study (GWAS) summary data from genetic consortia. Multivariable MR was then applied to evaluate the direct effect of reproductive behaviors on these psychiatric disorders whilst accounting for other reproductive factors at different life periods.

**Results:** Univariable MR analyses provide evidences that age at menarche, age at first sexual intercourse and age at first birth have effects on one (depression), seven (anxiety disorder, ADHD, bipolar disorder, bipolar disorder II, depression, PTSD and schizophrenia) and three psychiatric disorders (ADHD, depression and PTSD) (based on *p* < 7.14 × 10^−4^), respectively. However, after performing multivariable MR, only age at first sexual intercourse has direct effects on six psychiatric disorders (Depression, Attention deficit or hyperactivity disorder, Bipolar disorder, Posttraumatic stress disorder, Anxiety disorders and Anorexia Nervosa) when accounting for other reproductive behaviors with significant effects in univariable analyses.

**Conclusion:** Our findings suggest that reproductive behaviors predominantly exert their detrimental effects on psychiatric disorders and age at first sexual intercourse has direct effects on psychiatric disorders.

## 1 Background

Psychiatric disorders have become the major contributors to overall morbidity and disability across the globe [1]. Approximately 30% of individuals suffer from psychiatric disorders across the lifespan [2] and the causes of psychiatric disorders may be different at different stages of life. Since psychiatric disorder is one of the leading cause of disease burdens, it is necessary to identify potential risk factors for further prevention [3].

Female reproductive behaviors, including age at menarche, age at first sexual intercourse, age at first birth, age at last birth and age at menopause, have important implications in reproductive health and evolutionary fitness. Some of these reproductive behaviors have been identified as risk factors for various psychiatric disorders [4-6], such as depression, schizophrenia [7] and bipolar disorders [8]. Taking depression as an example, multiple studies in the past decade have examined the association between age of menarche and risk of depression in adolescent girls and adulthood [9-13]. For instance, a mendelian randomization analysis showed early menarche is associated with higher levels of depressive symptoms at 14 years old [11]. Other studies have implicated early age at first sexual intercourse and early age at first birth were risk factors of depression [14-17]. A recent study used the National Longitudinal Study of Adolescent Health dataset to show earlier age of first coitus was associated with depressive symptoms [14].

However, the potential role of a woman’s reproductive behaviors in the causal pathways between reproductive behaviors and psychiatric disorders from the perspective of the whole life-course remains unclear. Whether an individual can reverse the impact of age of early reproductive behaviors (such as menarche) on psychiatric disorders through lifestyle modifications is unclear, particularly as those who had smaller age at menarche tend to have smaller ages at other reproductive behaviors than normal people [18-22]. For example, a meta-analysis study showed that early menarche increases the risk of premature and early menopause by 80% [20]. This makes it challenging to distinguish whether reproductive behaviors in early life have independent and lasting effects on psychiatric disorders, or whether its effects are completely mediated by other reproductive behaviors in later years. If the latter is the case, then it is possible to avoid the potential adverse consequences of earlier reproductive behaviors by attaining a different reproductive lifestyle in the later years.

Mendelian randomization (MR) is an approach that can deal with these challenges by using genetic variants as instrumental variables (IV) to infer causality among correlated traits. A valid IV must satisfy the following three assumptions: 1) Relevance: IV is robustly related to the exposure; 2) Exchangeability: IV is independent of any confounders of the exposure and outcome relationship; and 3) Exclusion restriction: IV affects the outcome only through the exposure [23]. The advantage of MR is that it could eliminate the impact of confounding and reverse causality compared with traditional regression analysis [24-25]. In addition, multivariable MR is recently developed to determine whether an exposure affects the outcome under the condition of other exposures [26] which makes it possible to explore the causal pathways between the psychiatric disorder and reproductive behaviors.

In this MR study, we evaluated causal relationships between the five reproductive behaviors based on their temporal order, and explored whether genetically predicted reproductive behaviors have effects on twelve psychiatric disorders, using genome-wide association studies (GWAS) summary data of reproductive behaviors and psychiatric disorders on large sample populations that are publicly available. For psychiatric disorders with strong evidence of genetically predicted effects with multiple reproductive behaviors, we applied multivariable MR to examine evidences of direct or indirect effects by accounting for reproductive behaviors with significant effects.

## 2 Methods

### 2.1 Data sources

#### Reproductive factors

The reproductive factors investigated in this study includes age at menarche (AAM), age at fist sexual intercourse (AFS), age at first birth (AFB), age at last birth (ALB) and age at menopause (ANM). GWAS summary statistics for age of menarche [27] and menopause [28] were from ReproGen consortium, including 252,000 and 201,323 participants respectively. For AFS and AFB, we used the summary statistics from the GWAS meta-analysis of 36 cohorts by Day *et al*. [29] study, including 542,901 and 418,758 individuals respectively. GWAS summary statistics for ALB were from UK Biobank cohort [30] with the sample size of 170,248. These reproductive factors were restricted in European ancestry. Details for datasets are listed in Table 1. We selected SNPs at a GWAS threshold of statistical significance with *p* < 5 × 10^−8^ and linkage disequilibrium (LD) *r*^2^ < 0.001(10000*kb*). The F-statistics of all reproductive factors were larger than 10 (Table 2), which means that these IVs strongly predict the traits [23]. Thus, it is sufficient to generate a strong genetic instrument based on the selected SNPs.

**Table 1.** Datasets

| Traits | Consortium | Sample size<br>(cases/controls) | Year | Pubmed<br>ID |
| --- | --- | --- | --- | --- |
| menarche | ReproGen | 252000 | 2014 | 28436984 |
| Age at first sexual intercourse | - | 397338 | 2021 | 34211149 |
| Age at first birth | - | 542901 | 2021 | 34211149 |
| Age at last live birth | MRC-IEU | 170248 | 2018 | - |
| menopause | ReproGen | 201323 | 2015 | 34349265 |
| Autism spectrum disorder | PGC | 46351(18382/27969) | 2017 | 30804558 |
| Depression | PGC | 500199(170756/329443) | 2019 | 30718901 |
| Attention deficit or hyperactivity disorder | PGC | 21191(4945/16246) | 2018 | 29325848 |
| Obsessive compulsive disorder | PGC | 9725(2688/7037) | 2017 | 28761083 |
| Schizophrenia | PGC | 130644(53386/77258) | 2022 | 35396580 |
| Bipolar disorder (all) | PGC | 413466(41917/371549) | 2021 | 34002096 |
| Bipolar disorder (I) | PGC | 64802(25060/449978) | 2021 | 34002096 |
| Bipolar disorder (II) | PGC | 22560(6781/364075) | 2021 | 34002096 |
| Posttraumatic stress disorder | PGC | 174659(23212/151447) | 2019 | 31594949 |
| Anxiety disorders | PGC | 77096(33640/43456) | 2016 | 26754954 |
| Anorexia Nervosa | PGC | 14477(3495/10982) | 2017 | 28494655 |
| Tourette syndrome | PGC | 14307(4819/9488) | 2018 | 30818990 |

**Table 2.** F statistics of IVs for exposures

| Exposure | nSNPs | F statistics |
| --- | --- | --- |
| menarche | 190 | 67.549 |
| menopause | 198 | 111.759 |
| Age at first sexual intercourse | 194 | 46.822 |
| Age at first birth | 65 | 38.556 |
| Age at last live birth | 6 | 130.993 |

#### Psychiatric disorders

The GWAS summary data for psychiatric disorders were retrieved from the Psychiatric Genomes Consortium (PGC). We chose 12 related psychiatric disorders as the outcome, including anxiety disorders (33,640 cases/43,456 controls) [31], anorexia nervosa (14,477 samples) [32], attention deficit or hyperactivity disorder (4,945 cases/16,246 controls) [33], autism spectrum disorder (18,382 cases/27,969 controls) [34], bipolar disorder (41,917 cases/371,549 controls) and two BD subtypes: bipolar I disorder (25,060 cases/449,978 controls) and bipolar II disorder (6,781 cases/364,075 controls) [35], major depressive disorder (170,756 cases/329,443 controls) [36], obsessive compulsive disorder (2,688 cases/7,037 controls) [37], posttraumatic stress disorder (23,185 cases/151,309 controls) [38], schizophrenia (67,280 cases/86,912 controls) [39] and Tourette syndrome (55,386 cases/77,258 controls) [40]. Details for datasets are listed in Table 1. Ethical review and informed consent had been obtained in all of the original studies.

### 2.2 Statistical analyses

#### Genetic Correlation

We used high-definition likelihood (HDL) inference for genetic correlation analysis. Compared with LD score regression (LDSC), HDL makes full use of the information of the variance-covariance matrix of the Z-score in GWAS summary statistics [41]. The reference panel used in HDL is 1000 Genomes phase 3 European reference panel.

#### Univariable MR

To access the causal associations between reproductive factors and psychiatric disorders, we used “TwosampleMR” R package for MR analysis [42,43]. The inverse variance weighted (IVW) method [23,44] was conducted as our primary MR method to access the causal associations of reproductive behaviors in pairs, along with reproductive behaviors and psychiatric disorders. We applied a conservative Bonferroni correction (i.e. *p* < 0.05/70 = 7.14 × 10^−4^) as a strong evidence and a nominal threshold (i.e. *p* < 0.05) as a weak evidence of causal relationship. We supplemented MR-Egger regression [45], weighted median [46], weighted mode [47], MR-robust adjusted profile score (MR-RAPS) [48] and MR-Mix methods [49] as our sensitivity MR analyses, which make different assumptions based on different effectiveness of the SNPs. The results from multiple methods of MR are helpful to evaluate the robustness of our MR estimations. The intercept of MR-Egger regression shows the average pleiotropic effect among all used SNPs under the InSIDE (Instrument Strength Independent of Direct Effect) assumption and a non-zero intercept means there exists directional pleiotropy. Additionally, we used applied MR-PRESSO (Mendelian Randomization Pleiotropy RESidual Sum and Outlier) [50] to identify and remove potential outliers. Egger’s test is used for pleiotropy test. Heterogeneity tests include two methods: IVW and MR-Egger.

#### Multivariable MR

Multivariable MR based on IVW method was subsequently applied in a two-sample setting. This statistical method is suitable for considering the genetically predicted effects of multiple risk factors (e.g. age at menarche and age at menopause) on an outcome (e.g. depression) simultaneously. This enables us to estimate the direct effects of a reproductive behavior in early life (i.e. the effect of age at menarche after accounting for age at menopause) and indirect effects (i.e. the effect mediated by age at menopause) on a psychiatric disorder. We applied this model using all genetic variants for the reproductive factors, which have nominal significant (*p* < 0.05) causal effects on outcomes, after undertaking linkage disequilibrium clumping based on *r*^2^ < 0.001 to ensure independence of our instruments. The statistical analyses of multivariable MR were performed using R package “MVMR” [51].

## 3 Results

### 3.1 Genetic Correlation

Among all the genetic correlation analyses between reproductive factors and psychiatric disorders, there were 26 pairs of traits had significant genetic correlations (*r*_*g*_ *range*: |0.07 - 0.69|) under a conservative Bonferroni correction (*p* < 0.05/60 = 8.33 × 10^−4^) and another 9 pairs of traits had nominal significant genetic correlations under *p* < 0.05. Results are shown in Figure 1. Age of first sexual intercourse had the highest genetic correlations with all psychiatric disorders except autism spectrum disorder and Tourette syndrome, while age at menarche only had strong evidence of genetic correlation with depression. Among all psychiatric disorders, depression had evidence of genetic correlation with all reproductive factors (a weak evidence with menopause and strong evidence with other four traits), while there was null genetic correlation between Tourette syndrome and reproductive factors.

**Figure 1.**
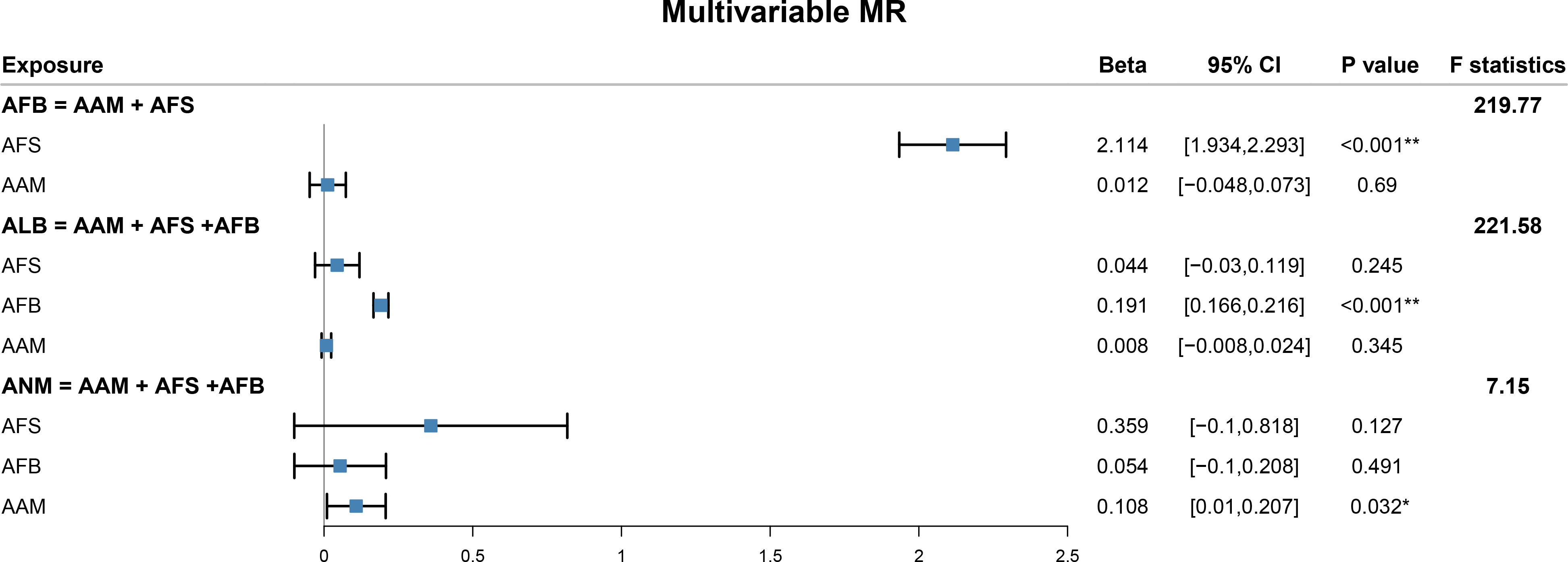
Genetic correlations between reproductive factors and psychiatric disorders.

### 3.2 Causal relationships among reproductive factors

#### Univariable MR

Results of univariable MR among reproductive factors are shown in Figure 2. The results of IVW method suggested that negative genetic correlation reflected causal effects of 190 SNPs determined later age at menarche (1 SD increase) on later AFS (b = 0.07; 95% confidence interval (CI) = 0.05,0.10), AFB (b = 0.17; CI = 0.09,0.25), ALB (b = 0.05; CI = 0.02,0.07) and ANM (b = 0.14; CI = 003,0.24) under Bonferroni-adjusted level of significance (*p* < 7.14 × 10^−4^). Results revealed that 194 SNPs determined later age at first sexual intercourse (1 SD increase) might lead to later AFB (b = 2.11; CI = 1.96,2.27), ALB (b = 0.43; CI = 0.37,0.48) and ANM (b = 0.49; CI = 0.19,0.79). For later age at first birth, its one SD increase led to later ALB (b = 0.19; CI = 0.17,0.20) and ANM (b = 0.15; CI = 0.05,0.26), using 65 SNPs as IVs. In addition, 6 SNPs determined age at last birth had no evidence of causal effect to ANM. These results were supported by weighted median, MR-RAPS methods (Figure S1) and MR PRESSO (Figure S2), suggesting the results were robust. In the results of ME PRESSO, we didn’t present the situation that outlier was not found. All the information of IVs were listed in Table S1. Egger’s test did not provide evidence that horizontal pleiotropy was driving these effects (Table S2). There is no heterogeneity in the Wald Ratio of all IVs (Table S3).

**Figure 2.**
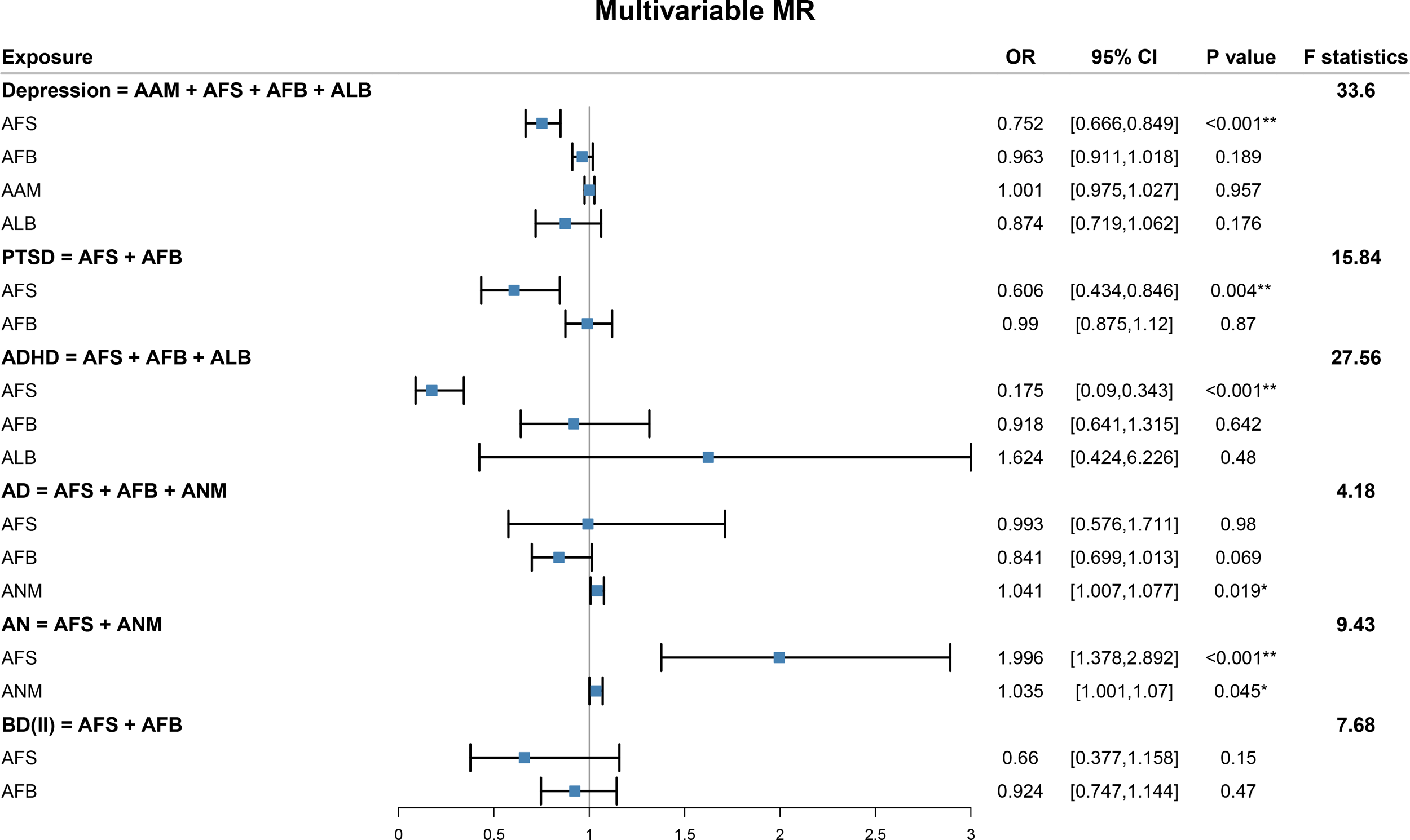
Univariable MR between reproductive factors and psychiatric disorders.

#### Multivariable MR

Results of multivariable MR were shown in Figure 3. Results suggested that only AFS had significant causal effect on AFB (b = 2.11; CI = 1.93,2.29), while AAM had no direct effects on AFB. After adjusting for AAM and AFS, AFB had significant direct effect on ALB (b = 0.19; CI = 0.17,0.27). We also found only AAM had positive causal effect on ANM (b = 0.11; CI = 0.01,0.21), while AFS and AFB had no direct effects on ANM.

**Figure 3.**
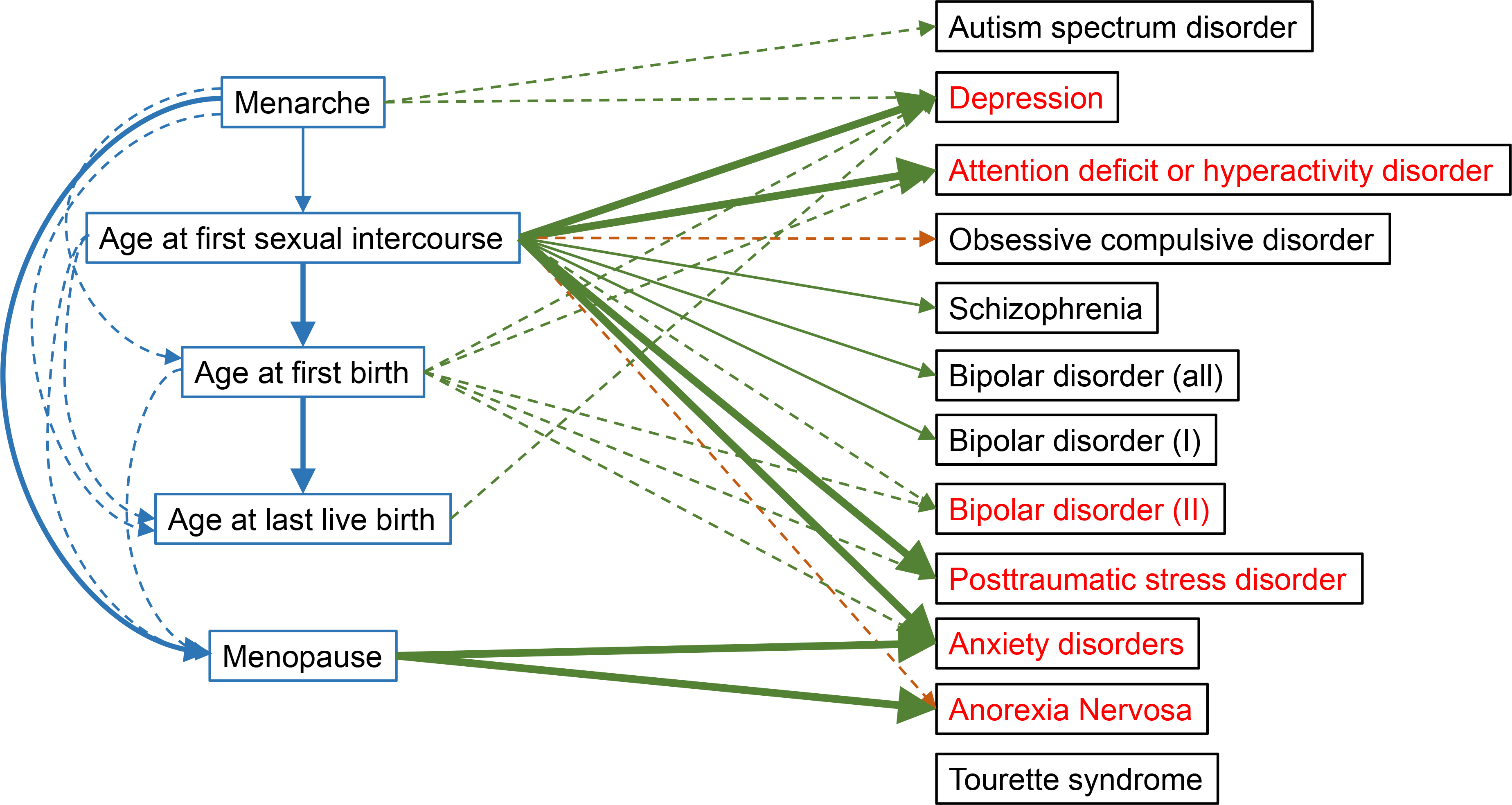
Multivariable MR among reproductive factors.

### 3.3 Causal relationships between reproductive factors and psychiatric disorders

#### Age at menarche

MR findings (Figure 2) from the primary analysis suggested that the negative genetic correlation reflects a causal relationship between later age at menarche (1 SD increase) and lower risk of depression (OR = 0.96; CI = 0.93,0.98) under Bonferroni-adjusted level of significance (*p* < 7.14 × 10^−4^). Results of weighted median and MR-RAPS methods were consistent with this result (Figure S3). Egger’s test did not provide evidence that horizontal pleiotropy (Table S4). Heterogeneity existed in the Wald Ratio of age at menarche and ADHD (Table S5). After removing outliers, MR PRESSO revealed that the later age at menarche reduced the risk of the ADHD (OR = 0.84; CI = 0.72,0.98) (Figure S4).

#### Age at first sexual intercourse

Results revealed that later age at first sexual intercourse (1 SD increase) may lead to lower risk of anxiety disorder (OR = 0.46; CI = 0.33,0.64), lower risk of ADHD (OR = 0.17; CI = 0.12,0.25), bipolar disorder (OR = 0.66; CI = 0.55,0.79), bipolar II disorder (OR = 0.58; CI = 0.43,0.79), depression (OR = 0.67; CI = 0.62,0.74), posttraumatic stress disorder (OR = 0.57; CI = 0.47,0.69) and schizophrenia (OR = 0.54; CI = 0.43,0.69). Although heterogeneity existed in several relationships (Table S5), these causal results were consistent with the results of weighted median, MR-RAPS methods (Figure S5) and MR PRESSO (Figure S6). Egger’s test did not provide evidence that horizontal pleiotropy (Table S4).

#### Age at first birth

We found that later age at first birth (1 SD increase) may lead to lower risk of ADHD (OR = 0.69; CI = 0.60,0.79), depression (OR = 0.87; CI = 0.84,0.90) and posttraumatic stress disorder (OR = 0.85; CI = 0.79,0.92). Results of weighted median and MR-RAPS methods were consistent with these results (Figure S7). Egger’s test did not provide evidence that horizontal pleiotropy (Table S4). There existed heterogeneity in the Wald Ratio of age at first birth and Bipolar disorder (Table S5). After removing outliers, MR PRESSO revealed that the later age at first birth reduced the risk of the Bipolar disorder (OR = 0.89; CI = 0.82,0.96) (Figure S8).

#### Age at last birth

MR results suggested that age at last birth had no evidence of causal effect to psychiatric disorders at Bonferroni-adjusted level of significance, but only nominal significant of later age at last birth (1 SD increase) lead to lower risk of depression (p = 0.003; OR = 0.65). Although heterogeneity existed in several relationships (Table S5), these causal results were consistent with the results of weighted median, MR-RAPS methods (Figure S9) and MR PRESSO (Figure S10). Egger’s test did not provide evidence that horizontal pleiotropy (Table S4).

#### Age at menopause

There was no evidence of causal effect of age at menopause on psychiatric disorders at Bonferroni-adjusted level of significance, but only nominal significant of later age at menopause (1 SD increase) lead to higher risk of anorexia nervosa (p = 0.02; OR = 1.04) and anxiety disorders (p = 0.02; OR = 1.04). Results of weighted median and MR-RAPS methods were consistent with these results (Figure S11). Egger’s test did not provide evidence that horizontal pleiotropy (Table S4). There existed heterogeneity in the Wald Ratio of age at menopause and Bipolar disorder (Table S5). After removing outliers, MR PRESSO revealed that the later age at menopause increased the risk of the Bipolar disorder (OR = 1.017; CI = 1.002,1.033) (Figure S12).

#### Multivariable MR

We used multivariable MR to evaluate the direct effects of multiple reproductive factors on the psychiatric disorders, resulted in the majority of effect estimates identified in the previous univariable analysis attenuating to include the null upon adjustment for other reproductive factors that had significant causal effect to the outcome (*p* < 0.05). Results were shown in Figure 4.

**Figure 4.**
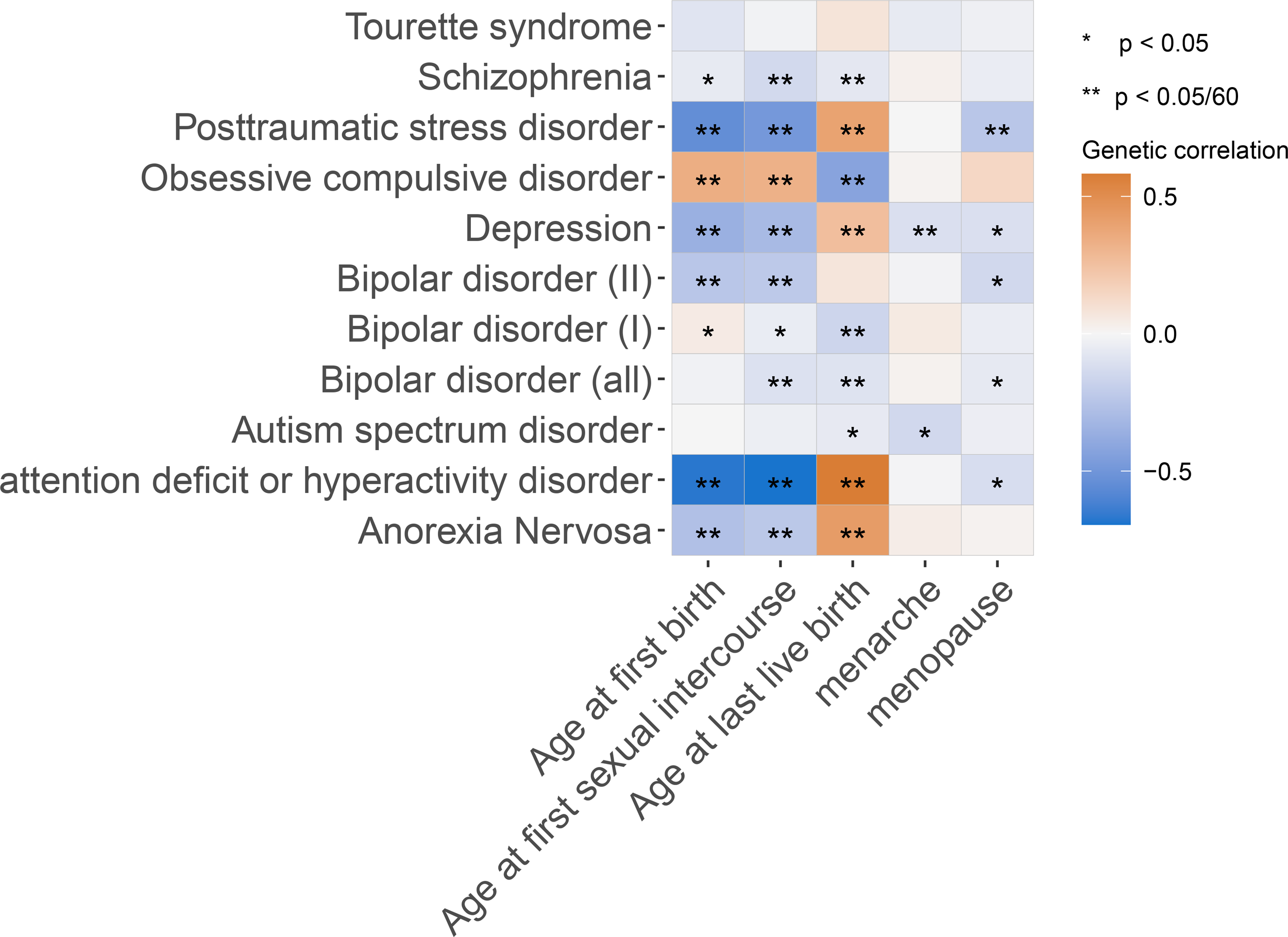
Multivariable MR between reproductive factors and psychiatric disorders.

We found that only AFS had significant causal effect to depression (OR = 0.75; CI = 0.67,0.85), while AAM, AFB and ALB had no direct effects to depression; only AFS had significant causal effect to PTSD (OR = 0.61; CI = 0.43,0.85), while AFB had no direct effect; only AFS had significant causal effect to ADHD (OR = 0.21; CI = 0.10,0.41), while AAM and AFB had no direct effects; only ANM had significant causal effect to anxiety disorders (OR = 1.04; CI = 1.00,1.08), while AFS and AFB had no direct effects; AFS and ANM both had significant direct effects to anorexia nervosa (OR = 2.00; CI = 1.38,2.90 for AFS and OR = 1.04; CI = 1.00,1.07 for ANM); neither reproductive factor had significant causal effect to bipolar II disorder.

This indicates that evidence of the total effects between AAM and ADHD and depression detected in the univariable analysis may due to the long-term effects of the earlier reproductive behavior across the life course (i.e. not just age at menarche). The evidence of the effect between AFS and anxiety disorder is likely totally mediated by ANM. The evidence of the effects between AFB and depression, PTSD, ADHD and anxiety disorder were likely confounded by AFS, suggesting there was no causal relations between AFB and these psychiatric disorders. The evidence of the effects between ALB and depression, ADHD and anxiety disorder were likely also confounded by AFS, suggesting there was no causal relations between AFB and these psychiatric disorders. Figure 5 is the causal graph that shows where we found evidence of an effect between reproductive behaviors and psychiatric disorders.

**Figure 5.**
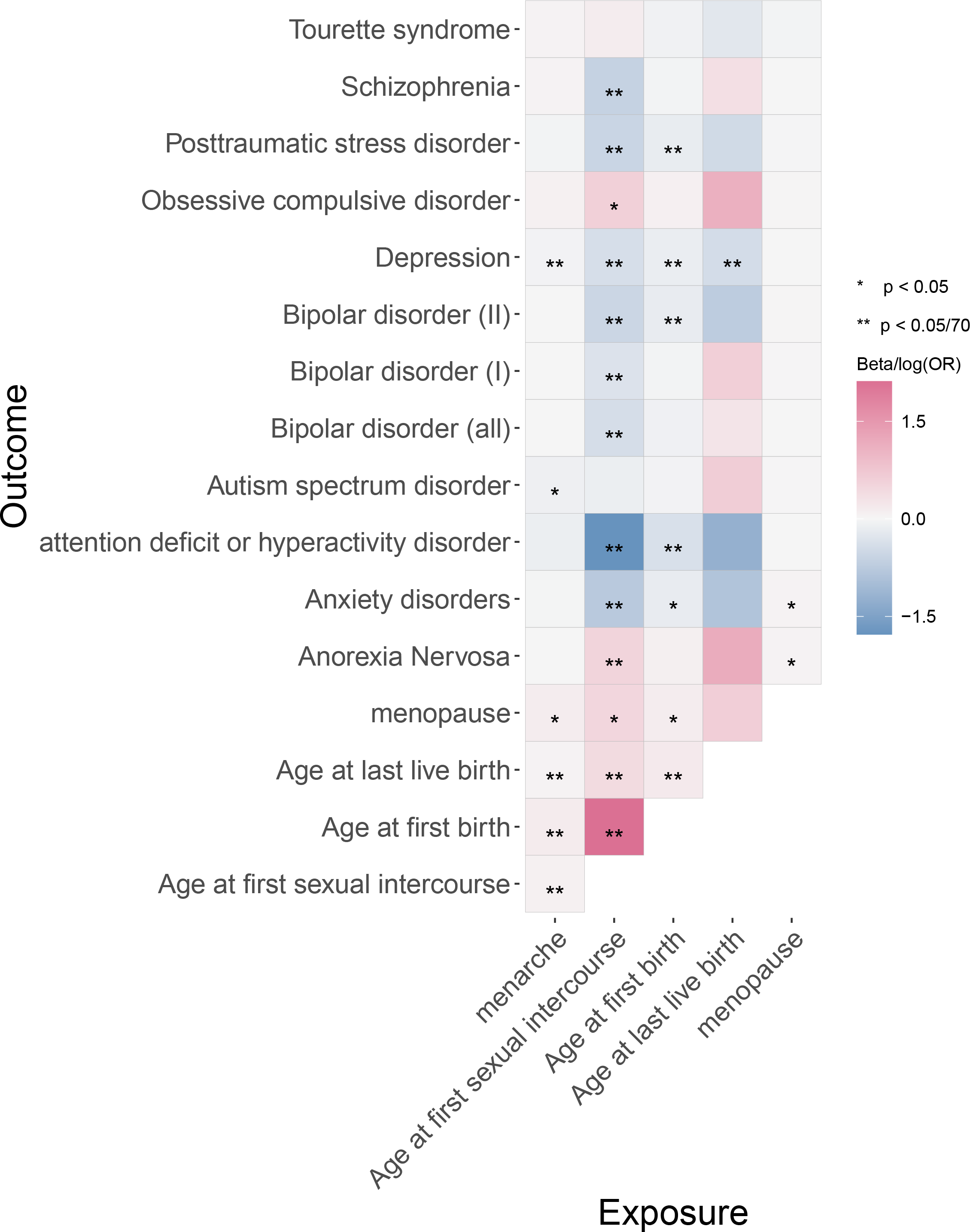
Causal relationships between reproductive factors and psychiatric disorders. The dashed lines represent the significant results in univariable MR but null significant results in multivariable MR or nominal significant results in univariable MR. The solid lines represent the significant results in both univariable and multivariable MR. The green lines represent the negative effects. The yellow and blue lines represent the positive effects. Psychiatric disorders with red color represent the diseases considered in multivariable MR analysis.

## 4 Discussion

We present evidence suggesting causal effects of several female reproductive behaviors on other reproductive traits. On the other hand, we show that later age of reproductive behaviors across the life course decreases the risk of psychiatric disorders, while later age at menopause increases the risk of anorexia nervosa and anxiety disorders. However, these effects estimation attenuated once accounting for other reproductive behaviors of different stages of life. On the one hand, these effects may be explained by the sustained impact of earlier sexual maturity in childhood and thus tend to remain so into adulthood. On the other hand, impact of common cause (i.e. reproductive factor during early life) is also important, and it has causal effects to both reproductive factors during adulthood and psychiatric disorders.

We corroborate the direct effects of age at menarche on AFS and age at menopause in previous studies [18,19,52,53], while showing the effects of age at menarche on AFB and ALB are totally mediated by AFS and AFB, respectively. Our study also supports positive causal links between AFS and AFB, AFB and ALB [21,53,54]. We show that earlier reproductive behaviors including age at menarche, AFS and AFB have effects on subsequent reproductive events through two main pathways: (i) the directed path from age at menarche to age at menopause; (ii) the pathway from age at menarche to ALB through AFS and AFB. Our findings suggest that for reproductive behaviors except age at menopause, an earlier reproductive behavior only have direct causal effect on the next reproductive behavior following the temporal order; and the age at menopause only affected by age at menarche. These findings oppose previous MR studies that supported causal effects between AFS and age at menopause or AFB and age at menopause [53,55-59].

For the causal relations between reproductive behaviors and psychiatric disorders, our study supports evidence for the earlier of age of reproductive behaviors in early life are causal factors in the rising risks of psychiatric disorders in previous studies [4-17]. However, the none effects of age at menarche, AFB and ALB after accounting for AFS using multivariable MR suggest that the effect of age at menarche on depression is totally mediated by AFS and the effects of AFB and ALB on psychiatric disorders are confounded by AFS. Our findings show that AFS is a direct risk factor on the pathway from reproductive behaviors to multiple psychiatric disorders, which has sustained impact on both reproductive behaviors and psychiatric disorders. The multivariable MR estimates for reproductive behaviors illustrate the importance of using a lifecourse MR approach to separate the effects of reproductive behaviors at separate stages in the life course [60-62].

One explanation for why earlier reproductive behaviors lead to earlier subsequent reproductive behaviors and psychiatric disorders is the life history theory. The core idea of this theory is to allocate limited resources between reproductive efforts and human growth, which can be divided into ‘fast’ and ‘slow’ life history strategies [63,64]. A ‘fast’ life history strategy makes more direct efforts to reproduce, such as earlier sexual intercourse which may lead to an earlier age of first birth, while people with a ‘slow’ life history strategy have later puberty and make a greater investment in a smaller number of offspring [63,64]. This theory is also supported by our study by showing that the women with an earlier age at menarche would have an earlier age at first sexual intercourse and earlier age at first birth [65]. On the other hand, women with a ‘fast’ life history may experience earlier age at menopause because of the huge resources on reproductive efforts in early life and the completeness of reproduction at a younger age [66,67], which is also supported by our results that earlier age at menarche leads to earlier age at menopause and earlier AFB leads to earlier ALB. Besides the age of reproductive behavior, life history theory can also explain the association between reproductive behaviors and psychiatric disorders. Life history theory explains the difference between ‘fast’ and ‘slow’ strategy as an adaptive response to an individual’s developmental environments and adverse childhood experiences have been shown to associate with earlier AFS, which has been identified as a risk factor for depression symptoms [68-70]. The ‘fast’ strategy in modern environments of earlier AFS is associated with teenage pregnancy and risk behaviors like violence, criminality, and substance abuse because of the little knowledge of reproductive health and these risk behaviors have been shown to be risk factors of multiple psychiatric disorders [71-73]. Our findings suggest that for preventing psychiatric disorders, it is important to control the age of first sexual intercourse rather than other reproductive behaviors like age at menarche by increasing sex education and parental monitoring.

Our research has various strengths and limitations. Firstly, we used data from large meta-analyzed sample of PGC consortia and ReproGen consortium, whose sample size is larger than any other studies before. On the other hand, the MR framework allowed us to avoid the effects of unmeasured confounders and reverse causation compared to more traditional epidemiology approaches [74] and the sensitive analysis of MR allowed us to evaluate the strong assumptions of MR. Furthermore, we used multivariable MR to investigate the causal pathways from five reproductive behaviors to twelve psychiatric disorders, which shows whether women with earlier menarche and first sexual intercourse could reduce the risk of psychiatric disorders by attaining a normal sexual lifestyle in later life. Conversely, one of the main limitations of our study is that all participants were of European descent. Therefore, it is not clear whether our findings are applicable to other ethnicities or populations of different ages, and further study needs to be discussed using samples from other ancestry groups or age groups [75-78]. In addition, the GWAS of the age of reproductive behaviors were derived from recall data with the possibility of recall bias, especially for reproductive behaviors during early life such as age at menarche and age at first sexual intercourse [75,76]. Another limitation of our study is that the ages of some important reproductive events are not included in our study such as the age of some reproductive diseases or the age at abortion.

## 5 Conclusion

In conclusion, our present lifecourse Mendelian randomization study provided evidence of causal relationships between five reproductive behaviors and twelve psychiatric disorders. In particular, we find strong evidence of the effects of reproductive behaviors on psychiatric disorders, and the importance of implementing preventative polices to control the age at first sexual intercourse in young age and its subsequent influence on the rising numbers of psychiatric disorders. This will help ease healthcare burdens and potentially improve the quality of life for individuals living with psychiatric disorders.

## Data Availability

All the GWAS summary data are publicly available. GWAS summary data for reproductive behaviors can be download at MR base (https://www.mrbase.org/). GWAS summary data for psychiatric disorders can be download at PGC consortium (https://pgc.unc.edu/).

https://pgc.unc.edu/

## Declarations

## Acknowledgement

We want to acknowledge the participants and investigators of the UK Biobank, MRC-IEU, ReproGen, PGC and MR base consortium.

## Conflict of interest disclosure

The authors declared no potential conflicts of interest with respect to the research, authorship and/or publication of this article.

## Funding statement

HL was supported by the National Natural Science Foundation of China (Grant 82003557) and Shandong Province Key R&D Program Project (2021SFGC0504). FX was supported by the National Natural Science Foundation of China (Grant 82173625).

## Ethics approval and patient consent statement

Ethical approval was not sought, because this study involved analysis of publicly available summary-level data from GWASs, and no individual-level data were used.

## Author contributions

FX, HL and TY conceived the study. YW, YY and SW contributed to data collection. YY, XL, YH, YG and YW contributed to the data analysis and sensitivity analyses. LH, FQ, QL and YF contributed to results collation and plot figures. YY, LH, XC, TY, HL and FX wrote and modified the manuscript. All authors reviewed and approved the final manuscript.

## Notes

### Competing Interest Statement

The authors have declared no competing interest.

