## Supplemental file for "Causal associations between female reproductive behaviors and psychiatric disorders: a lifecourse Mendelian randomization study"

Supplemental Figures

**Content**

Table S1. Instrumental variables in MR analyses

Table S2. Heterogeneity test of reproductive factors

Table S3. Egger’s test of reproductive factors

Table S4. Heterogeneity test of reproductive factors on psychiatric diseases

Table S5. Egger’s test of reproductive factors on psychiatric diseases

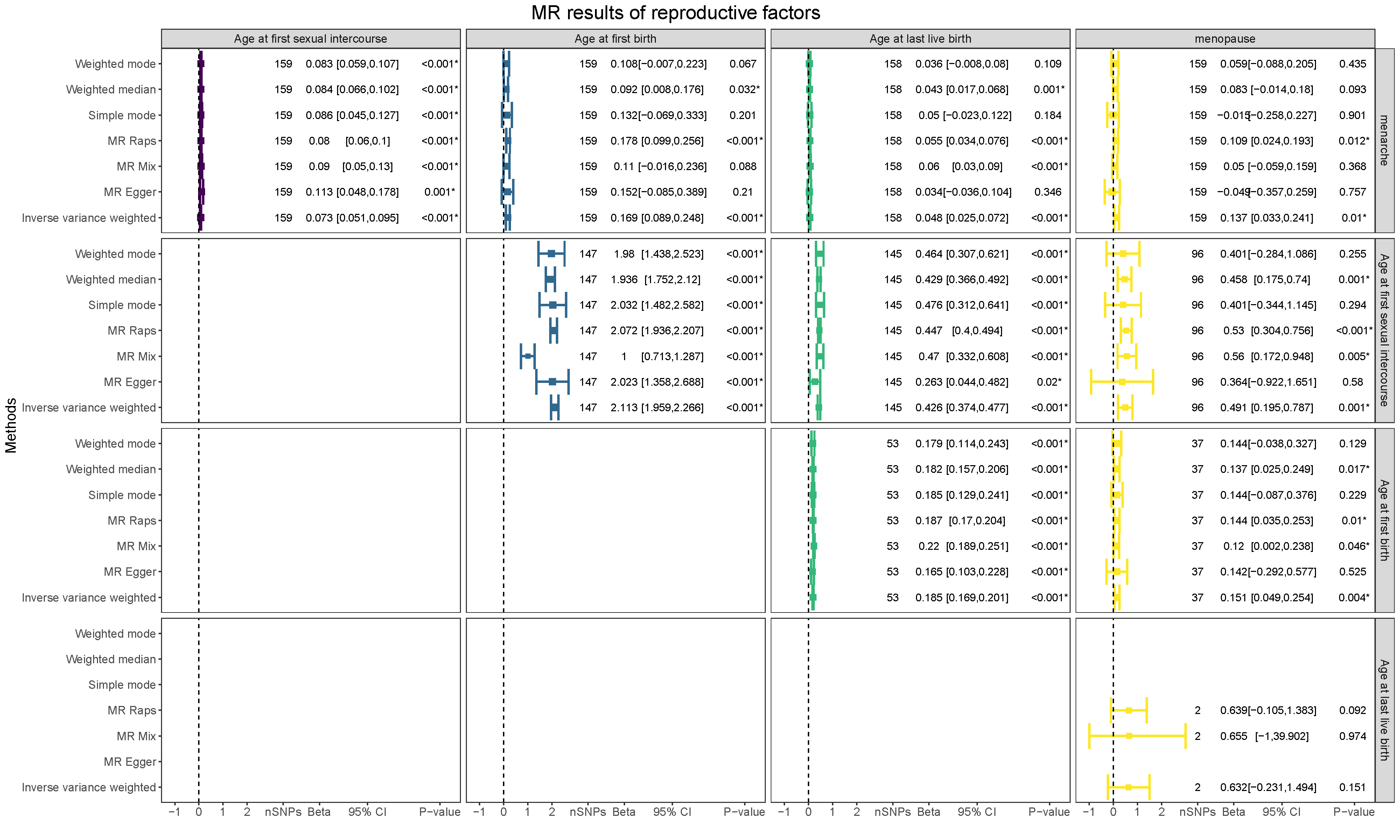

### Figure S1. MR results of reproductive factors

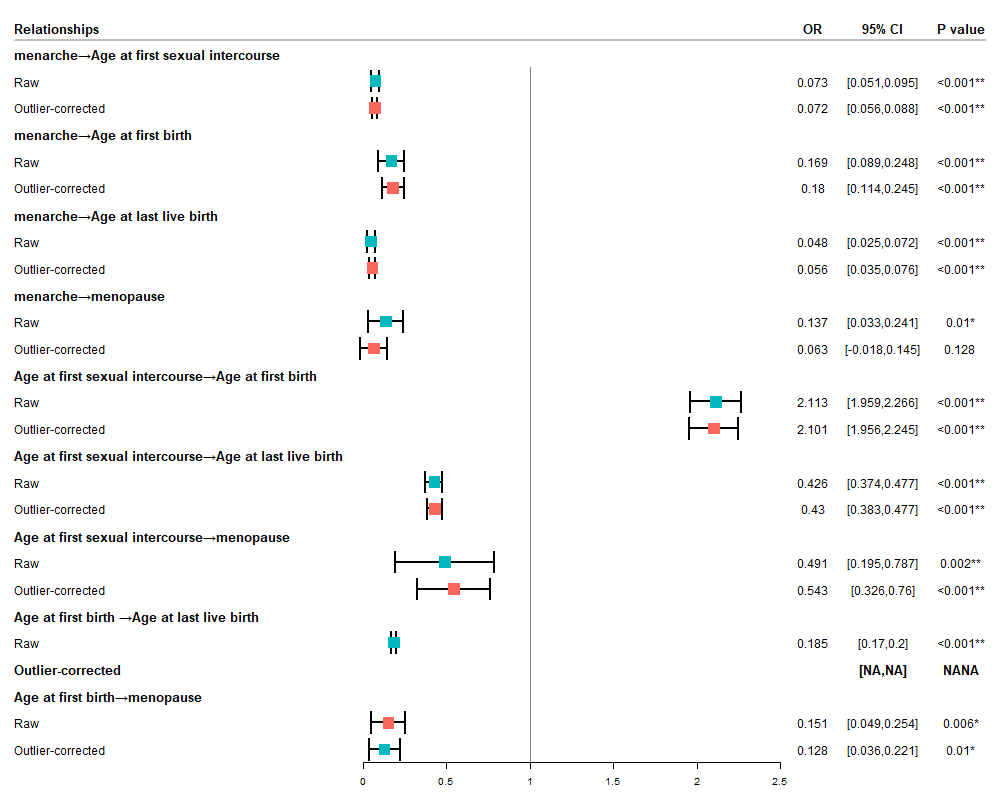

### Figure S2. MR PRESSO results of reproductive factors

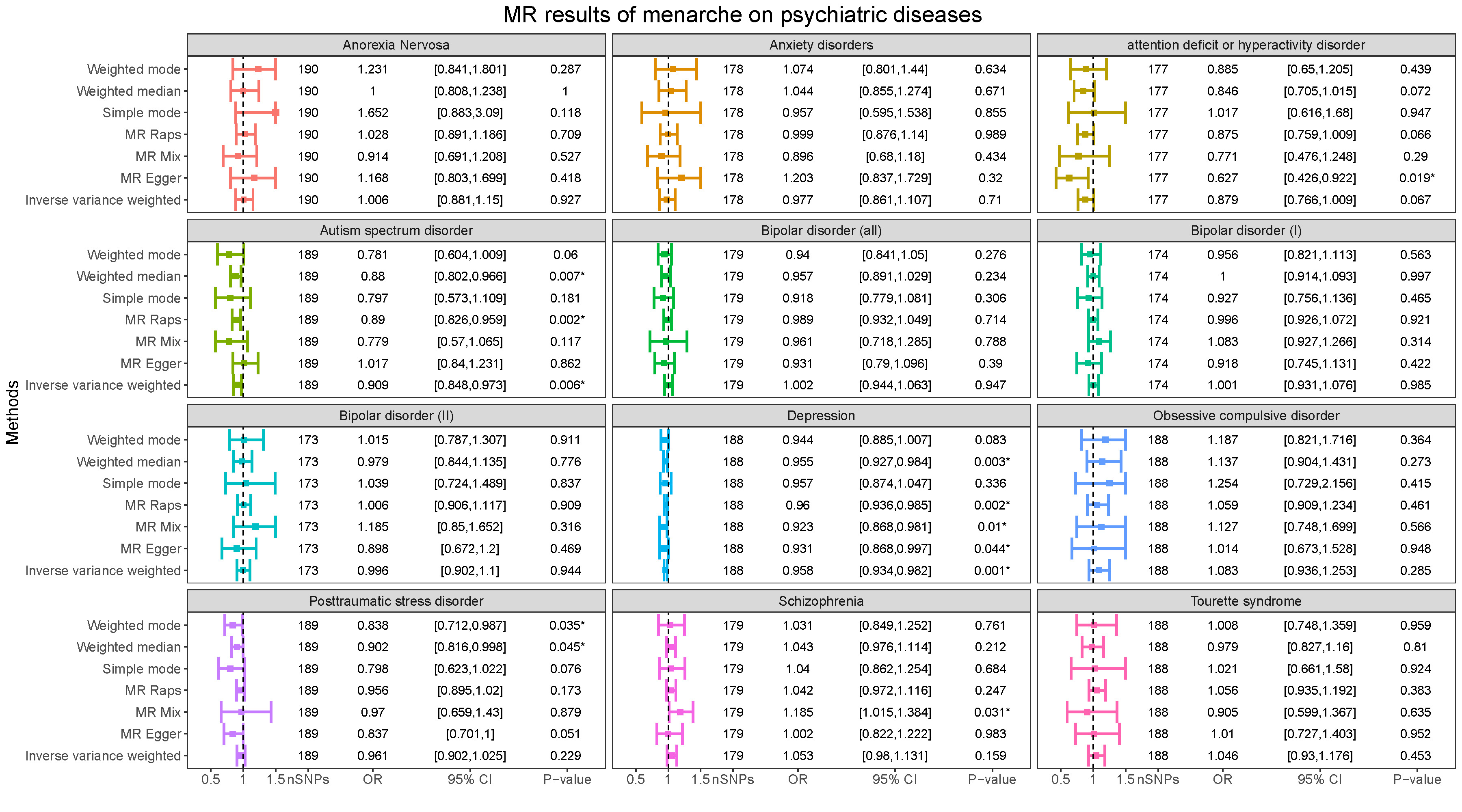

### Figure S3. MR results of age at menarche on psychiatric diseases

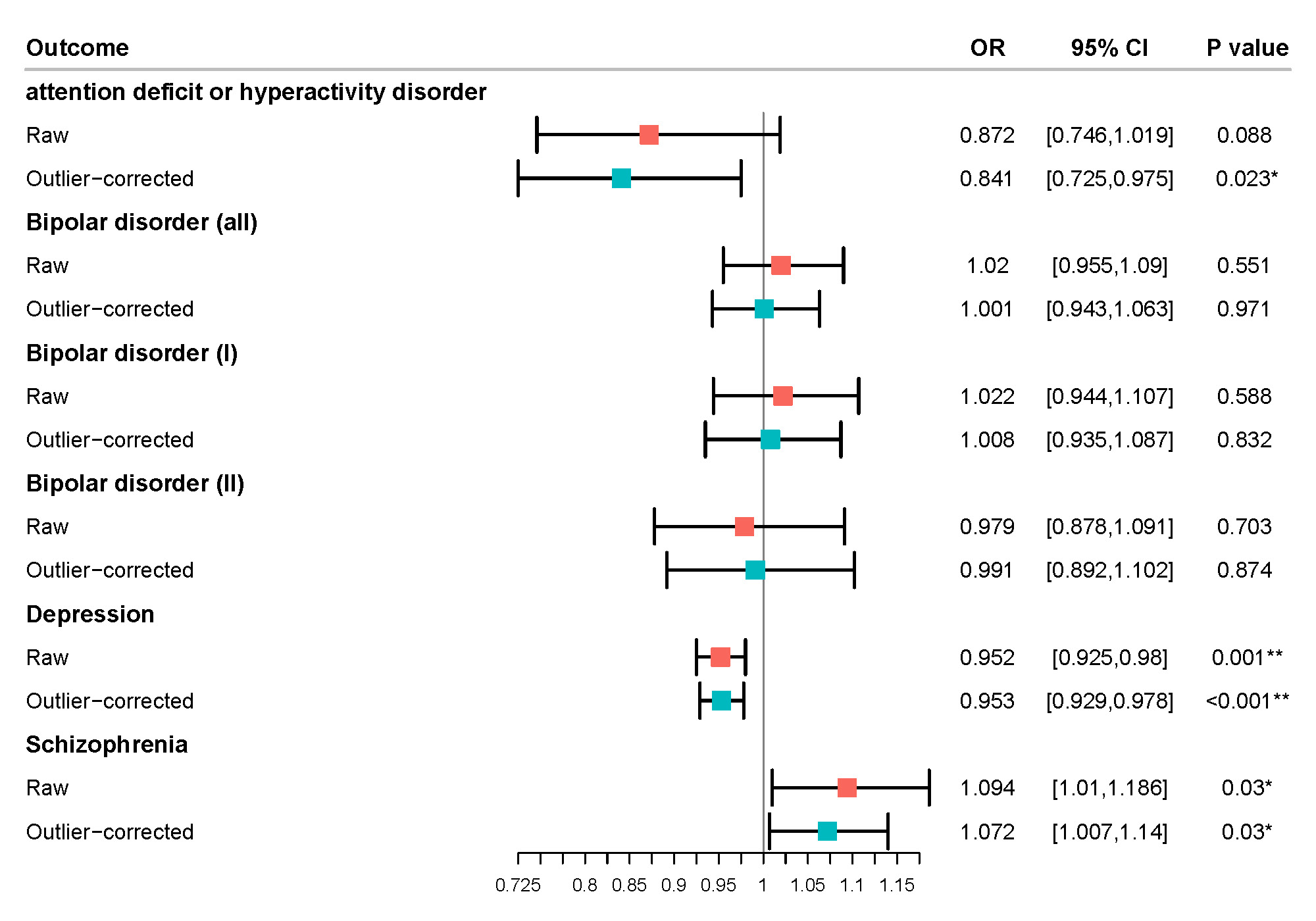

### Figure S4. MR PRESSO results of age at menarche on psychiatric diseases

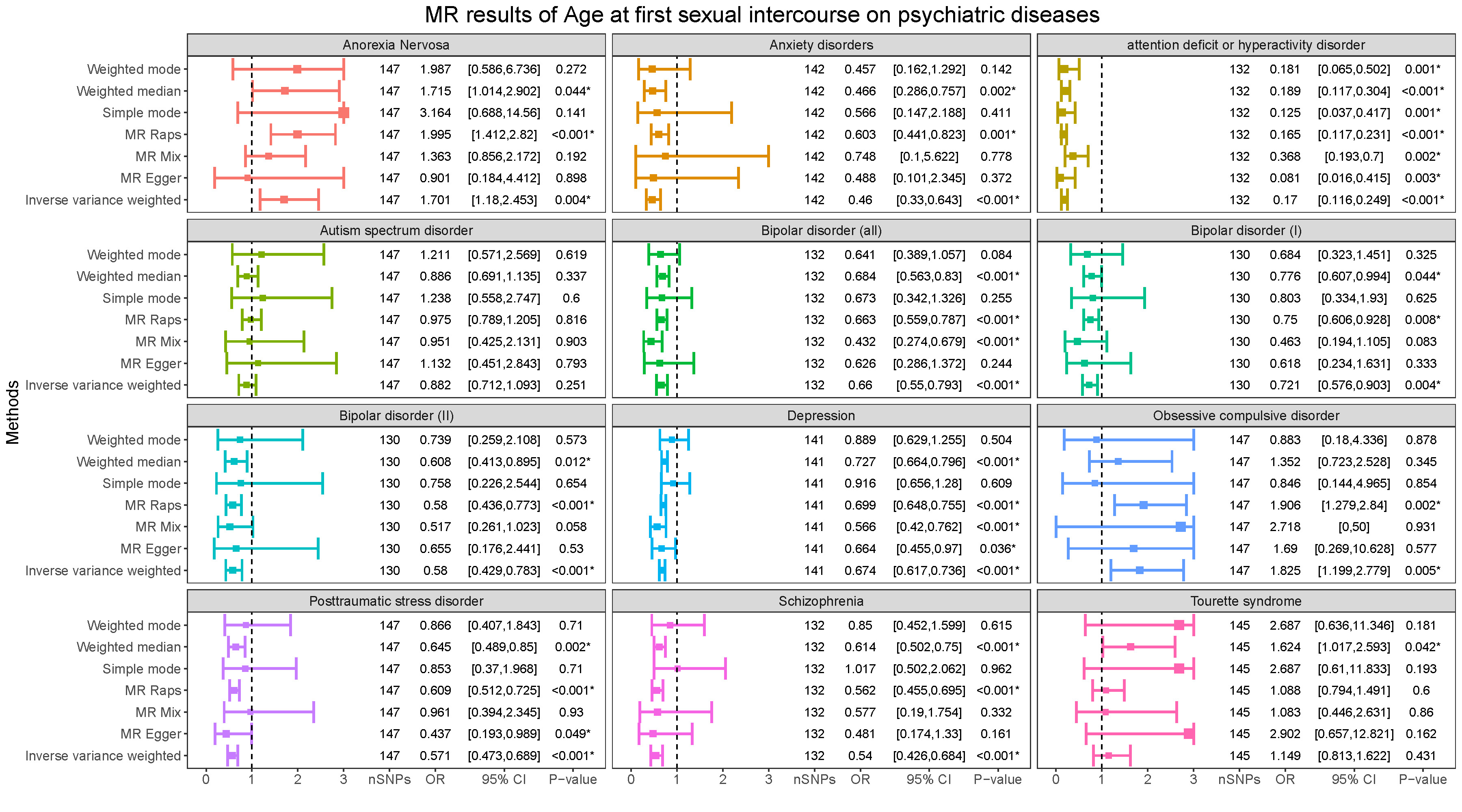

### Figure S5. MR results of age at first sexual intercourse on psychiatric diseases

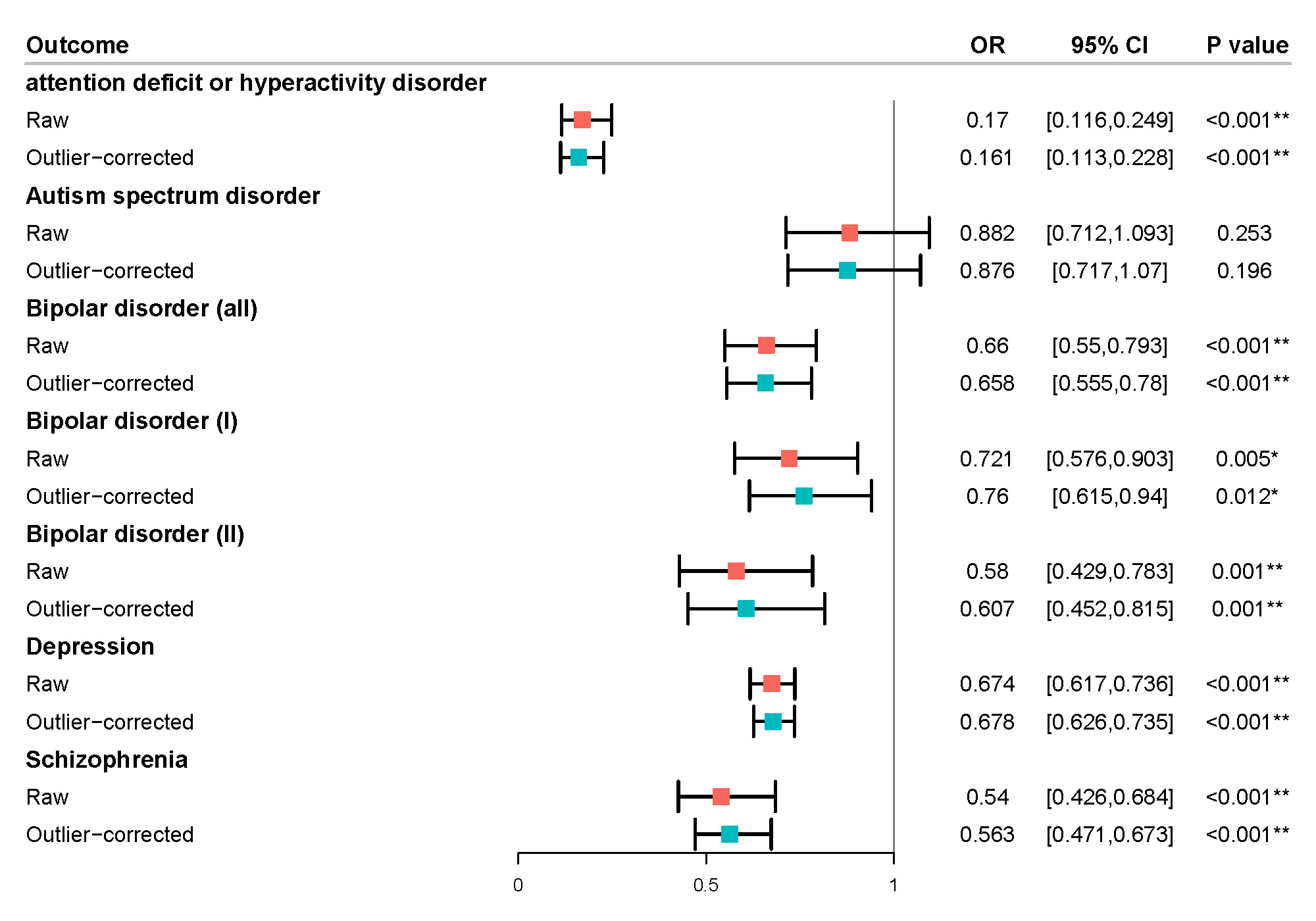

### Figure S6. MR PRESSO results of age at first sexual intercourse on psychiatric diseases

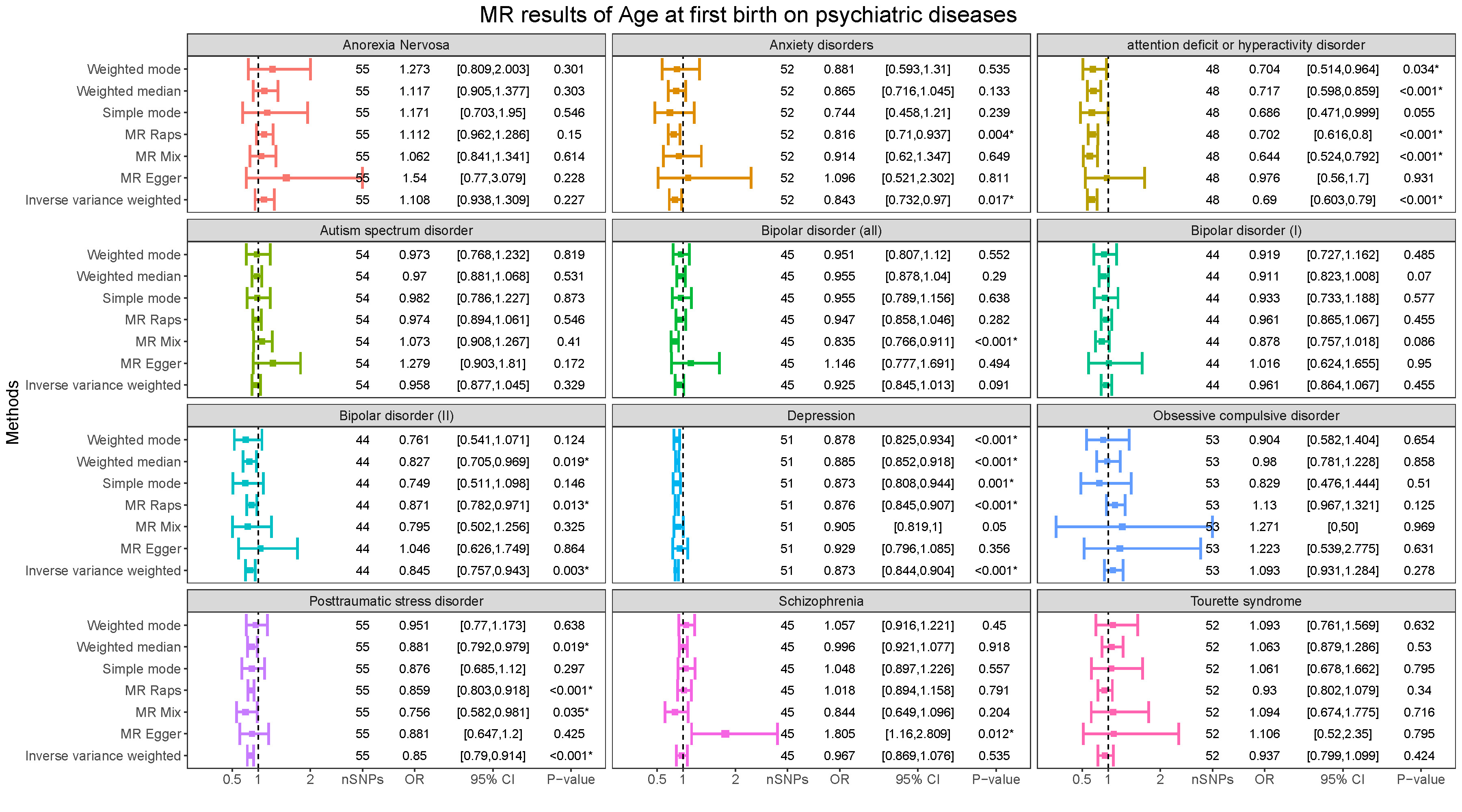

### Figure S7. MR results of age at first birth on psychiatric diseases

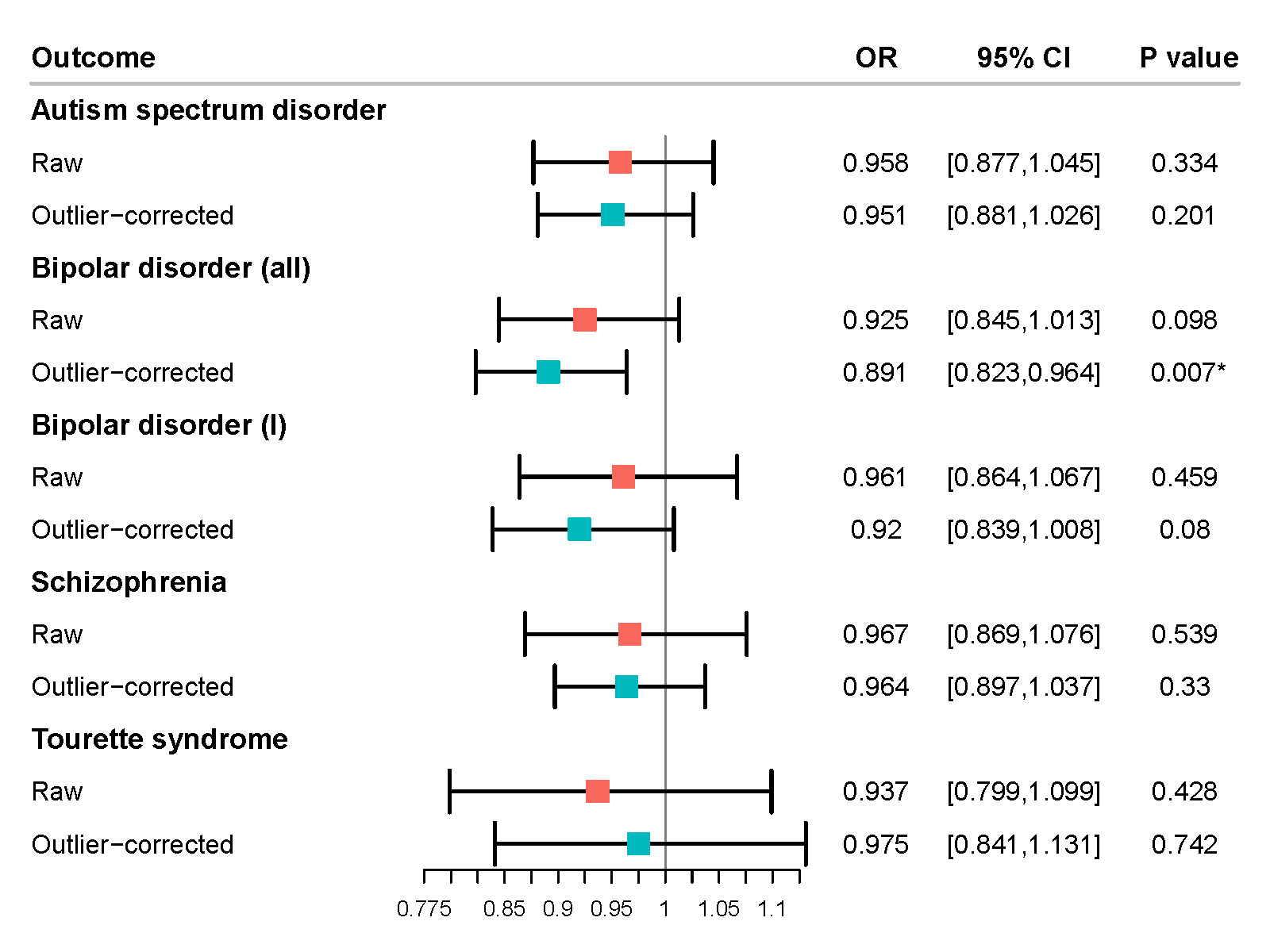

### Figure S8. MR PRESSO results of age at first birth on psychiatric diseases

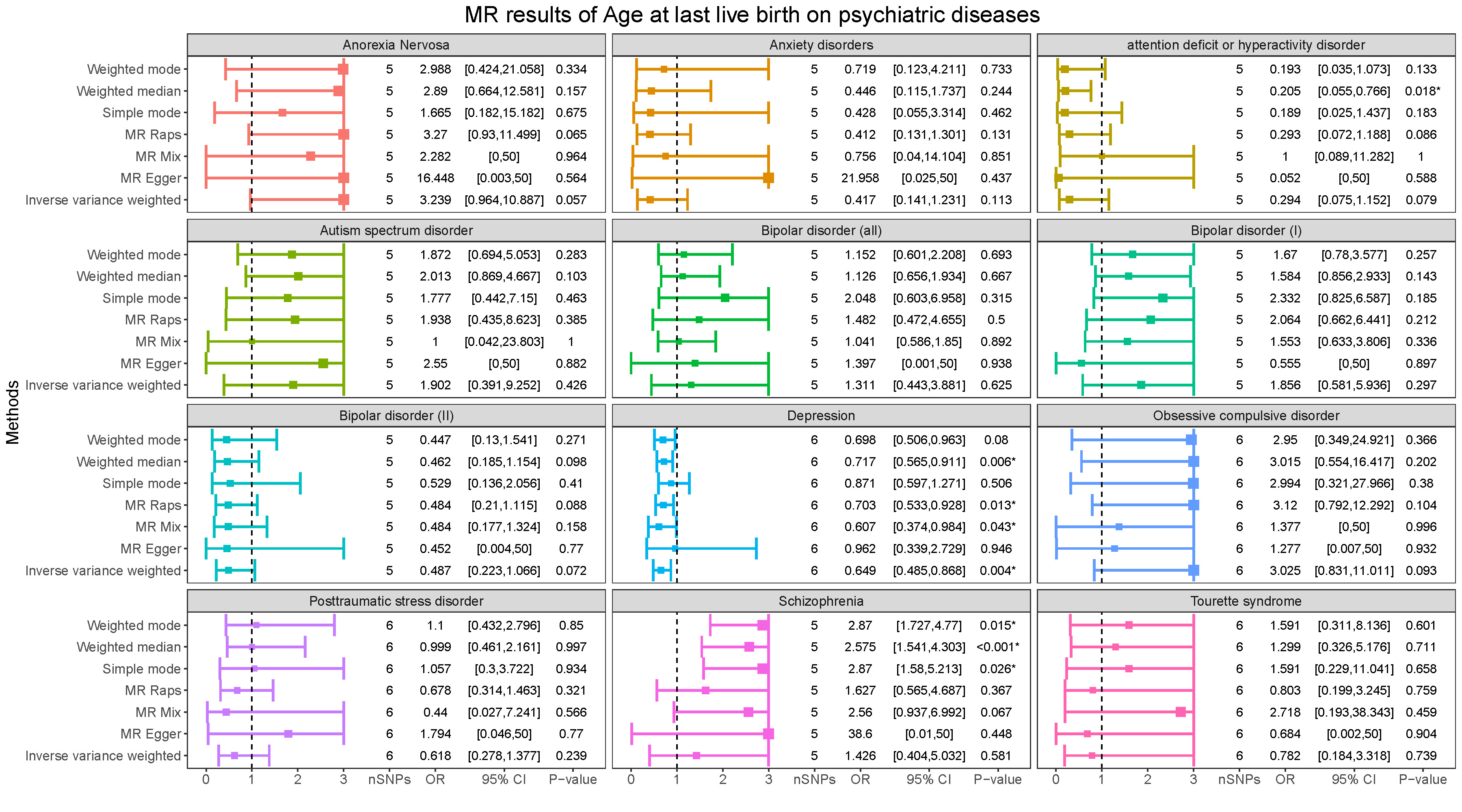

### Figure S9. MR results of age at last live birth on psychiatric diseases

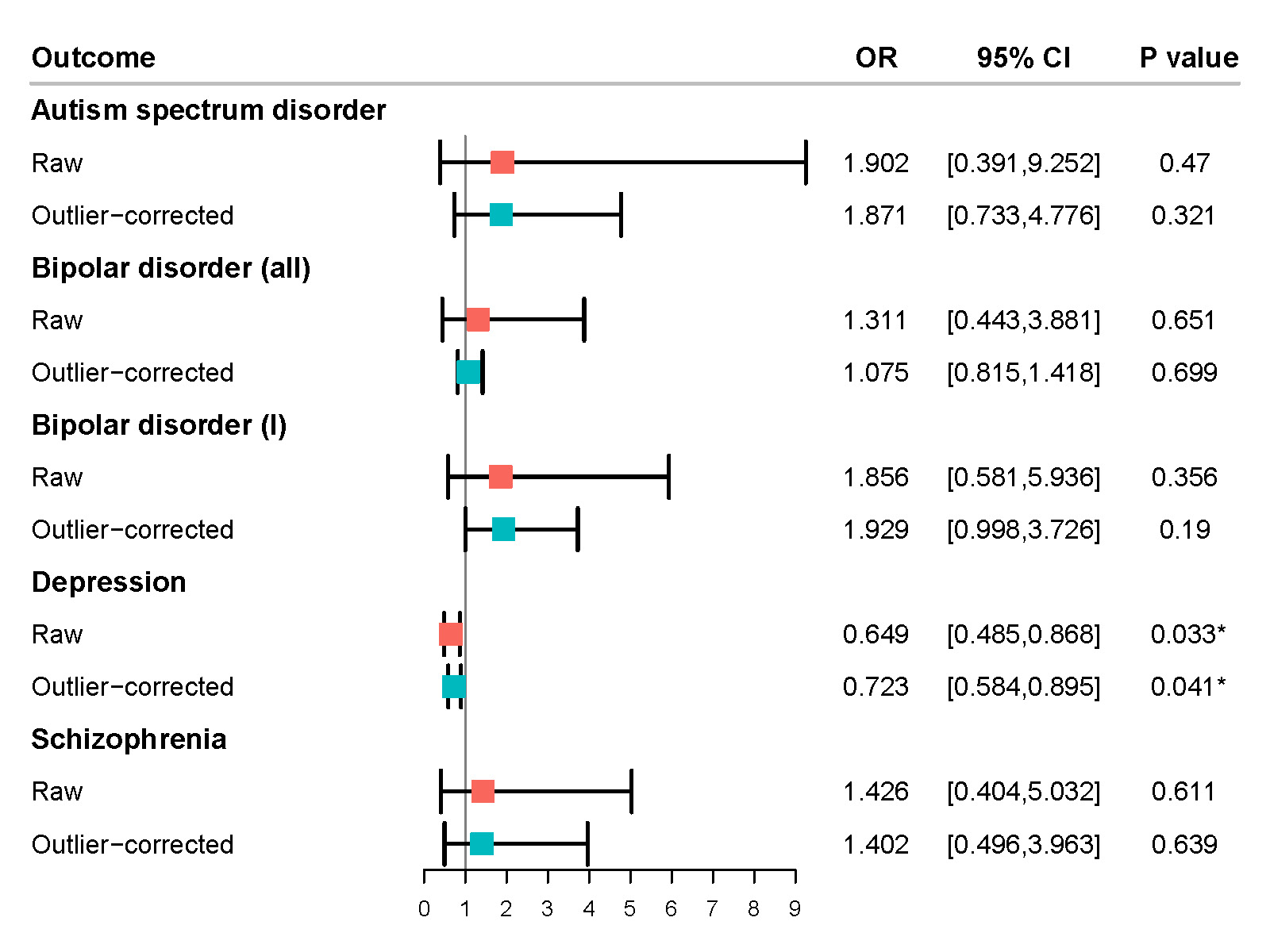

### Figure S10. MR PRESSO results of age at last live birth on psychiatric diseases

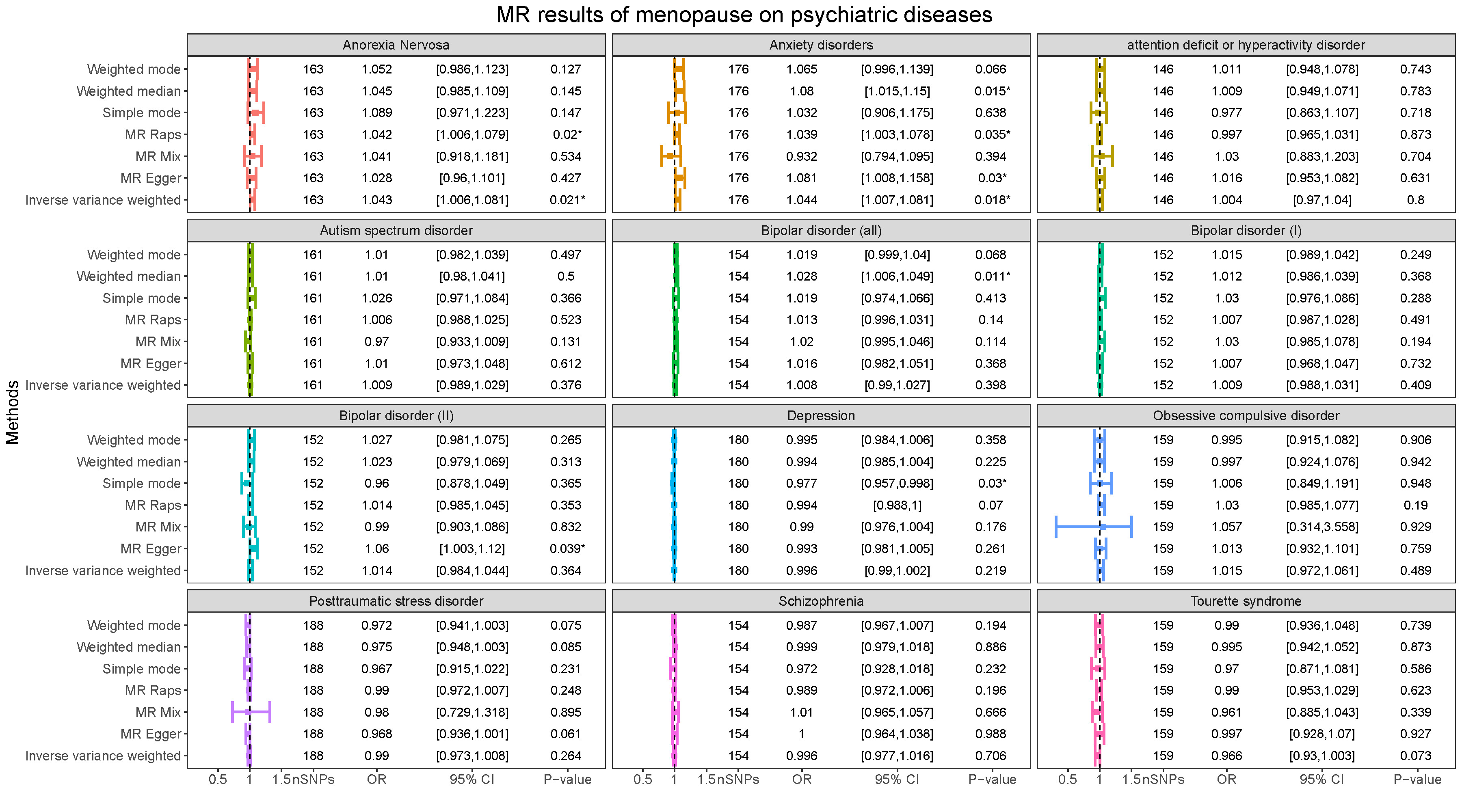

### Figure S11. MR results of age at menopause on psychiatric diseases

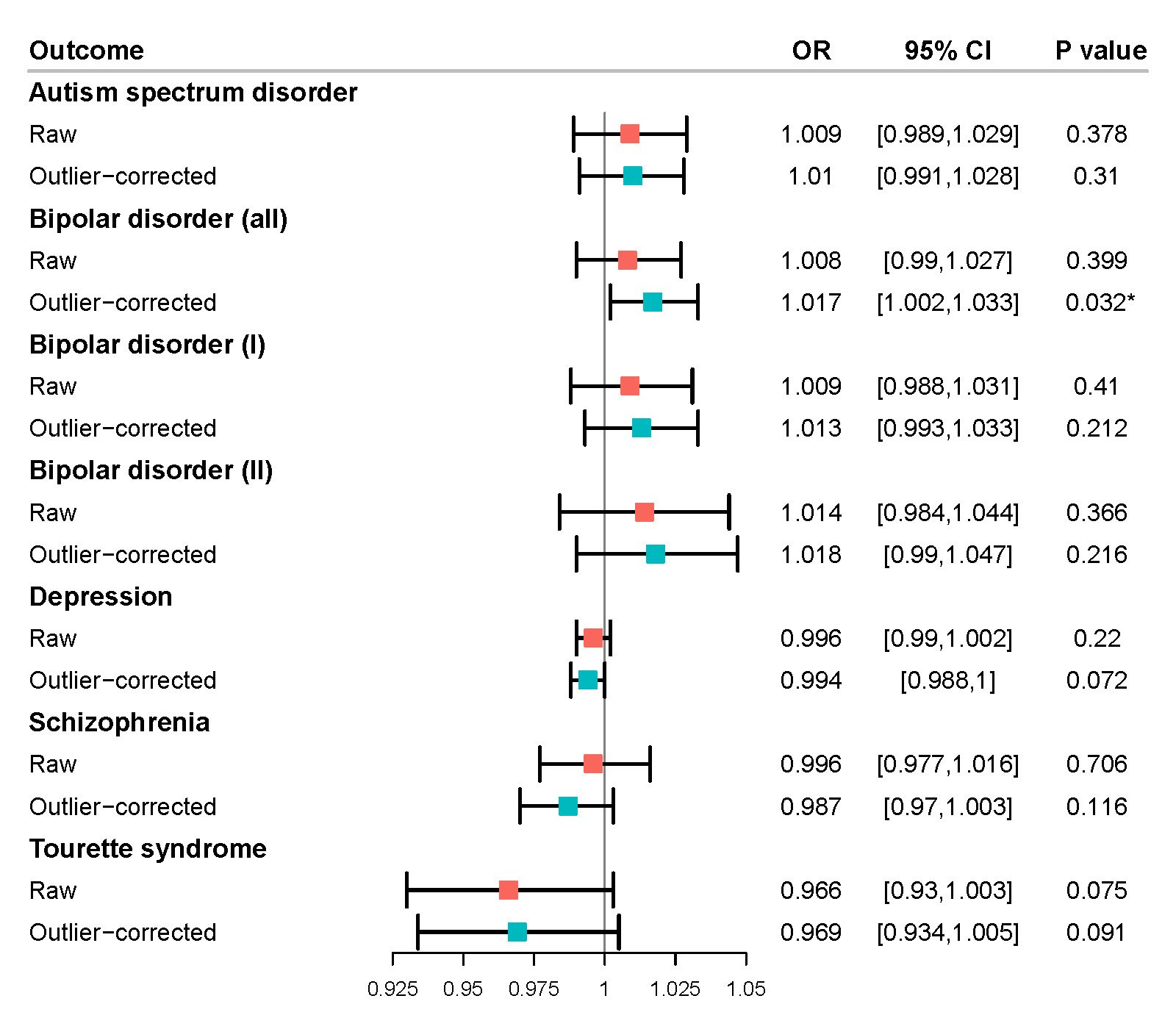

### Figure S12. MR PRESSO results of age at menopause on psychiatric diseases
